# The clinical outcomes and effectiveness of mHealth interventions for diabetes and hypertension: a systematic review and meta-analysis

**DOI:** 10.1101/2020.02.20.20025635

**Authors:** Yaqian Mao, Wei Lin, Junping Weng, Gang Chen

## Abstract

**Background:** With the development of technology, mobile health (mHealth) intervention has been proposed as a treatment strategy for chronic diseases that could improve the quality of chronic care and outcomes in some developed countries. However, the effectiveness of mHealth intervention in developing countries is not clear.

**Purpose:** A systematic review and meta-analysis was conducted to study the clinical outcomes and effectiveness of mHealth interventions for diabetes and hypertension in countries with different levels of economic development.

**Methods:** Pubmed, ResearchGate, Embase and Cochrane documents were searched by computer, and the retrieval period was from 2008 to June 2019. All studies were randomized controlled trials (RCTs) comparing mHealth treatments to other traditional treatments. Meta-analysis was conducted using stata software.

**Results:** 51 RCTs (N=13,054 participants) were eligible for this systematic review and meta-analysis. Compared with the usual care, the mHealth interventions yielded significant mean differences in clinical outcomes, and had a positive effect on countries at different levels of economic development. It is reassuring that we found mHealth interventions combined with human intelligence could significantly improve clinical outcomes more than mHealth interventions alone [WMD (95%Cl)=−6.75 (−9.98, −3.52)] VS [WMD (95%Cl)=−2.53 (−4.99, −0.07)]. The main secondary outcomes showed that mHealth interventions could also improve quality of life, satisfaction and self-efficacy, etc.

**Conclusion:** This review shown that mHealth interventions as a therapeutic strategy could improve the management of diabetes and hypertension in countries with different levels of economic development.

What is already known about this subject?

- With the development of technology, mobile health (mHealth) intervention has been proposed as a treatment strategy for chronic diseases that could improve the quality of chronic care in some developed countries.
- Nowdays, there are many mHealth products on the market, whether these mHealth products can improve the patient’s clinical results or improve the quality of life are still lack of research.

What are the new findings?

- mHealth interventions as a therapeutic strategy could improve the management of diabetes and hypertension in countries with different levels of economic development.
- mHealth interventions combined with human intelligence could significantly improve clinical outcomes more than mHealth interventions alone.

How might these results change the focus of research or clinical practice?

- mobile health (mHealth) intervention can as a treatment strategy for chronic diseases, especially in the undeveloped places.
- mHealth intervention treatments combined with special staff management (pharmacist, dietitian, specialist nurse and sports trainer) had more effective than mHealth interventions alone.

## Introduction

Diabetes mellitus (DM) and hypertension (HTN) are major modifiable risk factors for cardiac, cerebrovascular, and kidney diseases^[1]^, and are prevalent illnesses that co-occur in many patients. In the 2015 global risk factor assessment, high blood pressure, high blood sugar and smoking^[2]^ were the top three risk factors for increased disability, and sequelae caused by these factors, such as heart disease and stroke, were the leading causes of death^[3]^. DM and HTN are growing in epidemic proportions and disproportionately affects lower income, diverse countries. How to better manage chronic diseases has become the key to global health problems.

The prevalence of DM has been increasing worldwide in the past two decades, especially at a particularly fast pace in some developing countries, such as China and India^[4–7]^. China has a large burden of diabetes, According to a 2010 national survey, 11 % of adults in China had diabetes, affecting 109.6 million individuals^[6]^. What’s more, the prevalence of HTN in China is also high and increasing. According to a study from China^[8]^ that 33.6% or 335.8 million of the Chinese adult population had HTN in 2010, But only 3.9% patient were controlled to the currently recommended target of BP <140/90 mmHg. So develop some lower-cost and more effective methods for disease treatment and self-management are greatly needs in some less developed areas.

With the development of technology, mobile health (mHealth) management model has gradually entered the public life. More and more people have mHealth equipment, including lower socio-economic status groups^[9–11]^. The use of mHealth interventions may be an economical and effective method to provide disease self-management, and change the behavior of patients^[12–14]^, especially in patients with lower socioeconomic status.

Nowdays, there are many mHealth products on the market, whether these mHealth products can improve the patient’s clinical results or improve the quality of life are still lack of research. Some early literature reviews focused on assessing the practicability in different smart medical devices or mobile applications (Apps) ^[15–18]^in the management of patients with chronic diseases, while few reviews evaluated the effectiveness of existing mHealth devices as a health tool in the management of diabetes and hypertension. Furthermore, the efficacy and applicability of mHealth intervention have been confirmed by many clinical trials in developed countries, but few RCTs performed in China and other developing countries. In order to test whether mHealth interventions can improve the clinical outcome and effectiveness in areas with different levels of economic development, we carried out the systematic review and meta-analysis.

## Methods

### Search strategy and selection criteria

In order to identify studies that have investigated the effectiveness of mHealth intervention in disease (DM and HTN) management, we searched PubMed, ResearchGate, Embase and Cochrane for relevant articles, published between January 2008 to June 2019, because 2008 was the first consumer-oriented mobile application launched ^[15]^. All articles included in this study are in English.

The terms we used in the PubMed, ResearchGate, Embase, Cochrane search included “telemedicine and diabetes mellitus”, “telemedicine and hypertension”, “telemedicine and blood pressure,high”, “telemedicine and blood pressures,high”, “telemedicine and high blood pressure”, “telemedicine and high blood pressures”, “mobile health and diabetes mellitus”, “mobile health and hypertension”, “mobile health and blood pressure,high”, “mobile health and blood pressures,high”, “mobile health and high blood pressure”, “mobile health and high blood pressures”, “Health, mobile and diabetes mellitus”, “Health, mobile and hypertension”, “Health, mobile and blood pressure,high”, “Health, mobile and blood pressures,high”, “Health, mobile and high blood pressure”, “Health, mobile and high blood pressures”, “mHealth and diabetes mellitus”, “mHealth and hypertension”, “mHealth and blood pressure, high”, “mHealth and blood pressures, high”, “mHealth and high blood pressure”, “mHealth and high blood pressures”, “telehealth and diabetes mellitus”, “telehealth and hypertension”, “telehealth and blood pressure, high”, “telehealth and blood pressures, high”, “telehealth and high blood pressure”, “telehealth and high blood pressures”, “eHealth and diabetes mellitus”, “eHealth and blood pressure, high”, “eHealth and blood pressures, high”, “eHealth and high blood pressure”, “eHealth and high blood pressures”, The search was limited to studies involving randomized controlled trial, humanstudies, and publication in English.

### Study inclusion and exclusion criteria

Inclusion criteria that we used were, 1) Participants in studies were diabetic or/and hypertensive patient, 2) All subjects in the intervention group used mHealth intervention to conduct health management of diseases, 3) All the experimental methods were RCTs, 4) Experimental results should include observation objectives, such as the primary results (blood glucose, blood pressure), and the secondary results (self-efficacy, quality of life, satisfaction, etc.), and 5) the study was published in English.

Exclusion criteria included, 1) The full text is not available, 2) The experimental design does not meet the basic scientific requirements, 3) The study subjects were pregnant women, cancer patients and other non-target intervention groups, 4) The experimental group did not use mHealth devices for intervention or mHealth devices were not the main intervention measures, 5) The study results did not include target intervention results.

### Data extraction

First, We extracted the mean and standard deviation of the clinical indicators (HbA1c, FBG, SBP, DBP) at the end of the intervention, to evaluate the difference between mHealth intervention treatment and traditional treatment. Second, we assessed the clinical outcomes (HbA1c, SBP) at countries with different levels of economic development that using the mHealth intervention, and evaluated the difference of combination therapy compare with mHealth alone. Last, we analyed the impacts of using mHealth in self-efficacy, satisfaction and healthy behaviours, etc. Two coauthors (Y. Mao, W. Lin) and a research (J. Weng) assistant extracted information from identified studies that met inclusion criteria, including study design, subjects, nature of the intervention, inclusion of control groups, etc. and key research results. Extracted information was reviewed by other coauthors to verify accuracy.

### Statistical analysis

The methodological quality of the selected studies was assessed by using the Jadad scale ^[19]^, which has been recognized as a useful tool for evaluation of RCT quality ^[20]^. Jadad scores range from 0 (very poor) to 5 (rigorous)^[21]^.

We used Stata sofeware for all statistical analyses. Heterogeneity among studies was measured with the *I*^2^, the magnitude of heterogeneity was divided into low (*I*^2^< 25%), moderate (*I*^2^ ≥50%), significant (*I*^2^≥75%). When there was no significant heterogeneity in the study (*I*^2^<50%), we use a fixed-effect model to pool the data. When heterogeneity was more than moderate in the study (*I*^2^≥50%), we use a random-effects model. Mean differences (MDs) and corresponding 95% CIs were calculated when studies had the same units or used same measurements. A sensitivity analysis was performed to examine the cause of heterogeneity.

We assessed the possibility of publication bias by constructing a funnel plot. We assessed funnel plot asymmetry using Begg and Egger tests, and defined significant publication bias as a *p* value <0·1, the Begg and Egger checks are completed with the metabias command. The trim-and-fill analysis was used to estimate the effect of publication bias on the interpretation of the results, which was completed by the metatrim command.

## Results

### Main characteristics of the studies

We identified 1747 studies, of which 51 (with data for participants) were included in our analysis (See **Figure 1**), a total of 13,054 subjects were represented in the studies.

**Figure 1.**
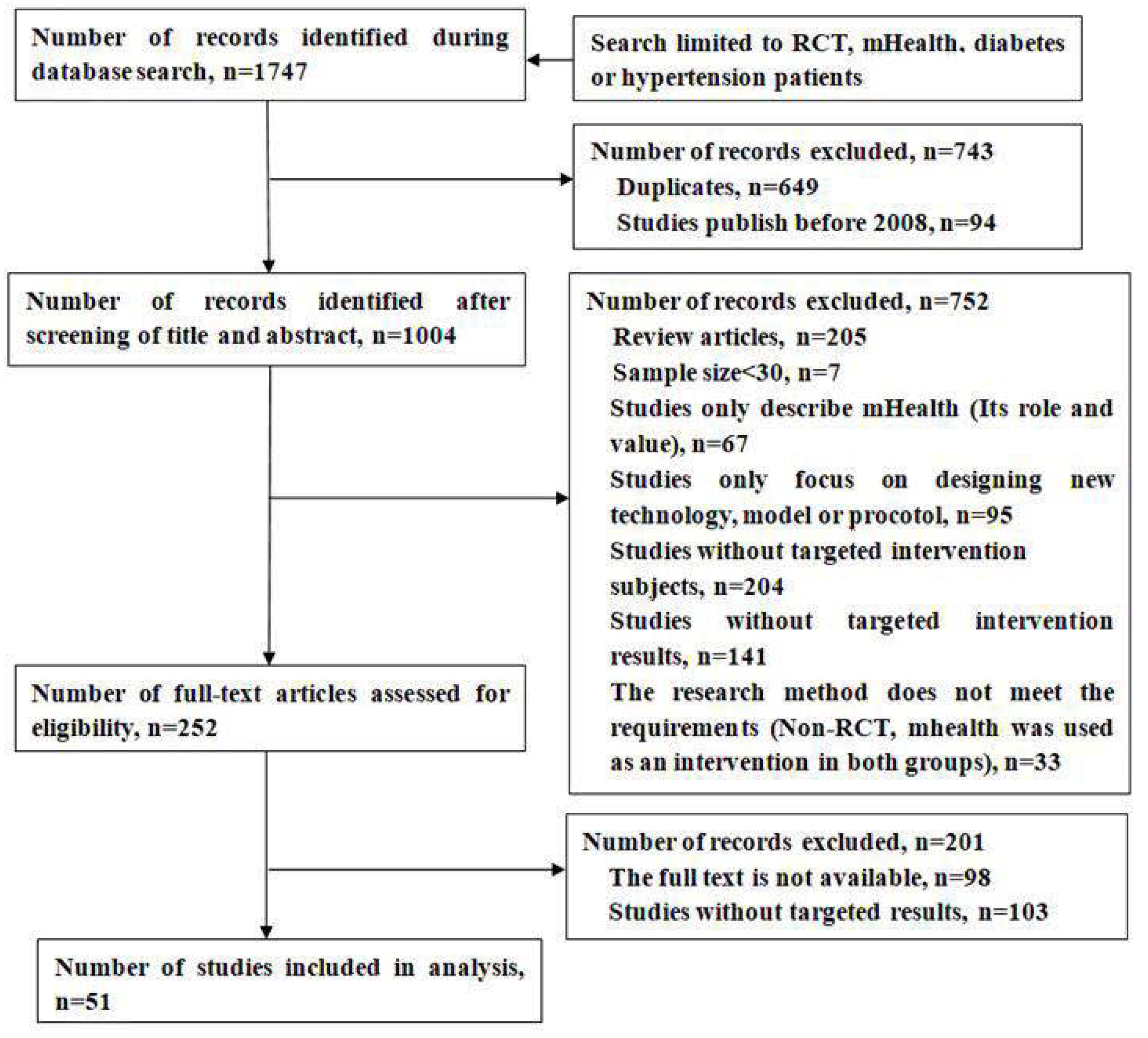
Flow chart of the literature search and study selection procedures. mHealth, mobile health;RCT, randomized controlled trial.

**Table 1** and **Table 2** shown the main characteristics of the 51 selected studies. Of these, 36 studies (70.59%) were conducted in developed countries, 15 studies (29.41%) were conducted in developing countries. All of the above study were RCTs.

**Table 1.**
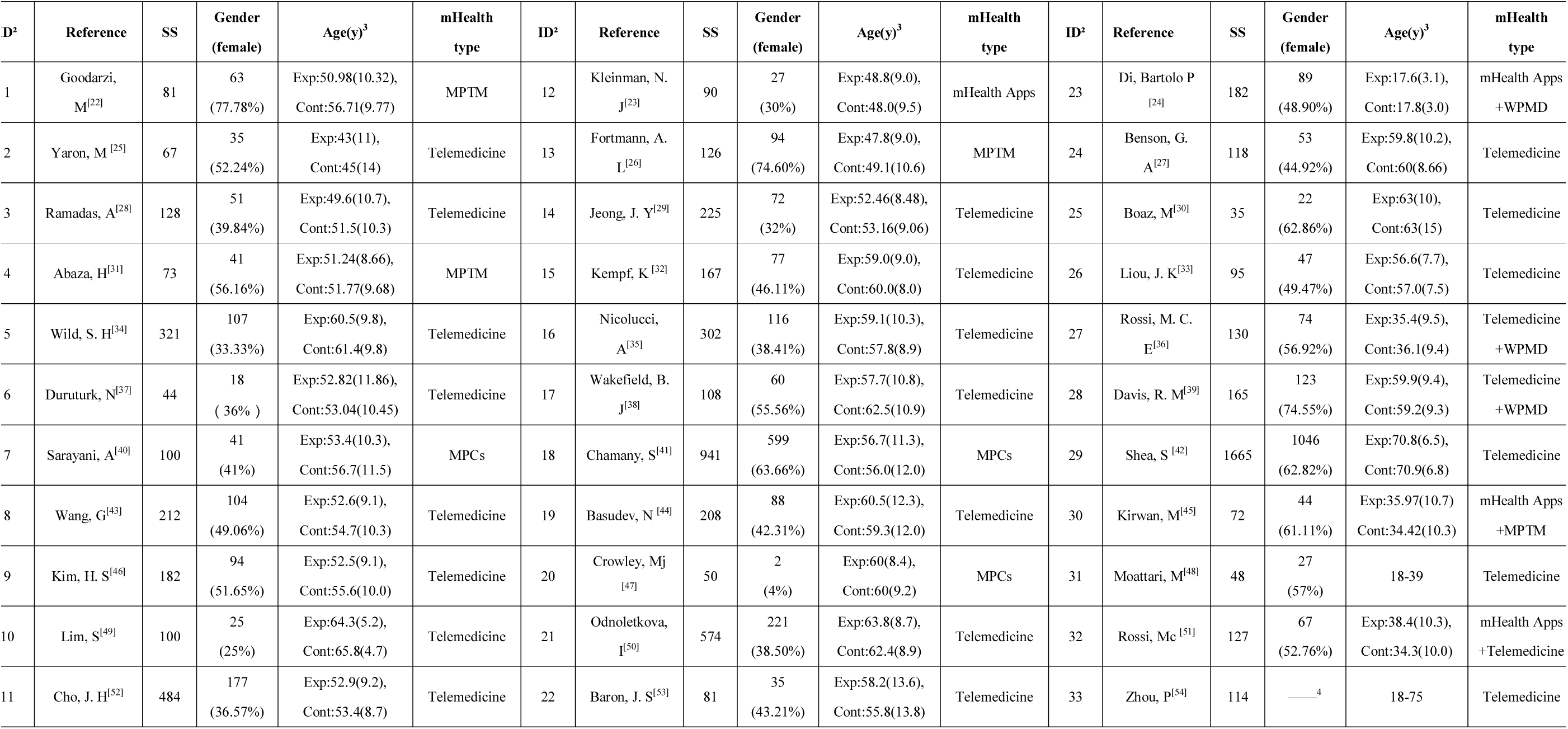
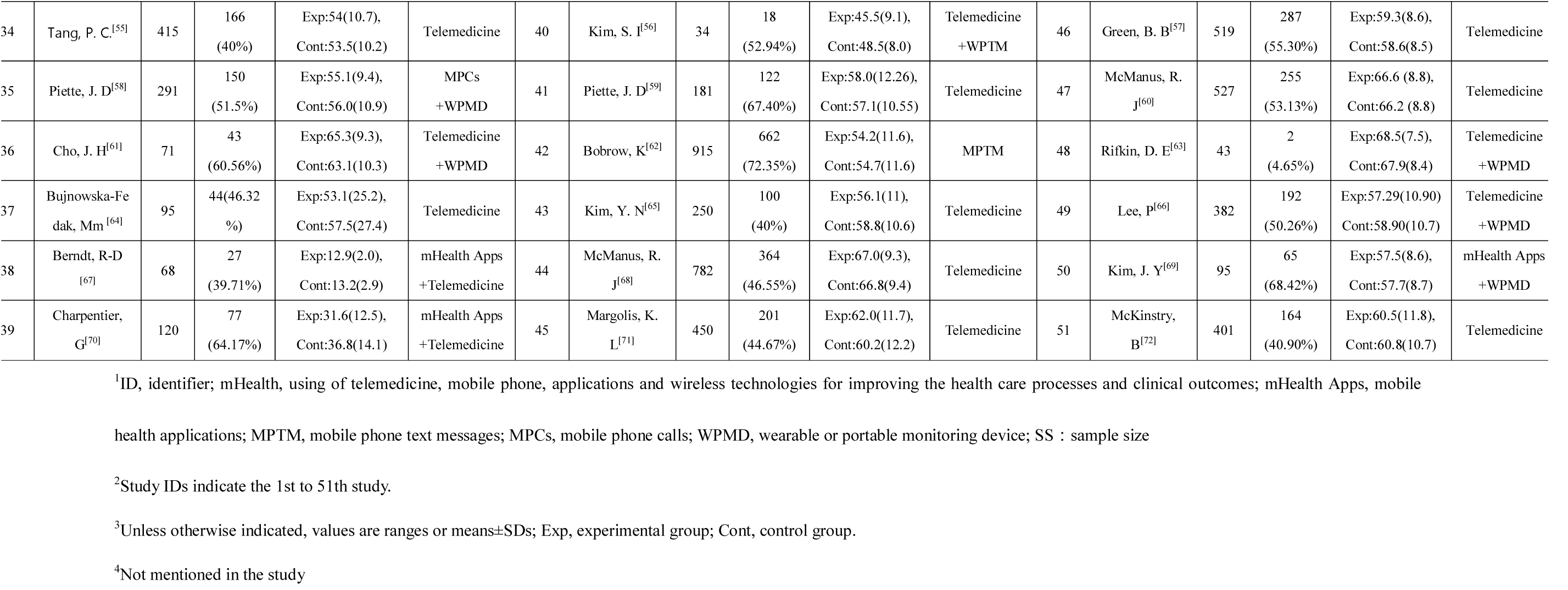
Summary of characteristics of 51 studies that examined mHealth interventions for hypertension and diabetes treatment and management^1^

**Table 2.** Summary of characteristics of 51 studies that examined mHealth interventions^1^ for hypertension and diabetes treatment and management

| Category | Number of studies(n, %) | Study ID<sup>2</sup> |
| --- | --- | --- |
| **Country/setting** |  |  |
| **Developed country** |  |  |
| Unite state | 13 (25.5%) | 13, 17, 18, 20, 24, 28, 29, 34, 35, 45, 46, 48, 50 |
| England | 6(11.8%) | 5, 19, 22, 44, 47, 51 |
| Korea | 6 (11.8%) | 10, 11, 14, 36, 40, 43 |
| Italy | 4 (7.8%) | 16, 23, 27, 32 |
| Germany | 2(3.9%) | 15, 38 |
| Israel | 2(3.9%) | 2, 25 |
| Australia | 1(2.0 %) | 30 |
| Belgium | 1(2.0%) | 26 |
| France | 1(2.0%) | 44 |
| **Developing country** |  |  |
| China | 5(9.8%) | 8, 9, 26, 33, 49 |
| Iran | 3 (5.9%) | 1, 7, 31 |
| Egypt | 1(2.0%) | 4 |
| India | 1(2.0%) | 12 |
| Honduras and Mexico | 1(2.0%) | 41 |
| Malaysia | 1(2.0%) | 3 |
| Turkey | 1(2.0%) | 6 |
| Poland | 1(2.0%) | 37 |
| South Africa | 1(2.0%) | 42 |
| **Intervention time/duration** |  |  |
| ≤3 mo | 11 (21.6%) | 1, 4, 6, 7, 15, 17, 31, 33, 36, 38, 41 |
| 3-6mo | 23(45.1%) | 3, 8-14, 20, 21, 23, 25, 26, 27, 30, 32, 37 39, 43, 48-51 |
| >6mo | 17(33.3%) | 2, 5, 16, 18, 19, 22, 24, 28, 29, 34, 35, 40, 42, 44-47 |
| **Sample size** |  |  |
| <100 | 17(33.3%) | 1, 2, 4, 6, 12, 20, 22, 25, 26, 30, 31, 36, 37, 38, 40 , 48, 50 |
| 100-500 | 27(52.9%) | 3, 5, 7-11, 13-17, 19, 23, 24, 27, 28, 32-35, 39, 41, 43, 45, 49, 51 |
| >500 | 7(13.7%) | 18, 21, 29, 42, 44, 46, 47 |
| **Targeted patient<sup>3</sup>** |  |  |
| T1DM | 7(13.7%) | 2, 23, 27, 30, 32, 38, 39 |
| T2DM | 28 (54.9%) | 1, 3-16, 19-21, 24, 26, 28, 29, 33-37, 40 |
| T1DM & T2DM combined | 4(7.8%) | 18, 22, 25, 31 |
| T2DM & HTN combined | 1(2.0%) | 17 |
| HTN | 11(21.6%) | 41-51 |

<sup>1</sup> mHealth <sup>[5]</sup>, using of telemedicine, mobile phone, applications and wireless technologies for improving the health care processes and clinical outcomes; MPTM, mobile phone text messages; MPCs, mobile phone calls; mHealth Apps, mobile health applications, WPMD, wearable or portable monitoring device, Telemedicine, distance therapy, management, education, rehabilitation, monitoring, and follow-up, etc
<sup>2</sup> Study ID, the related references for each study.
<sup>3</sup>T1DM, type 1 diabetes mellitus; T2DM, type 2 diabetes mellitus; HTN, hypertension

The study period ranged from 2008 to 2019, and the total sample size of the selected studies varied from 34 to 1,665 subjects. Each study included both male and female subjects. Intervention durations ranged widely from only 1 month to 5 years, most studies (23, 45.1%) had an intervention time between 3 and 6 months, 11 studies (21.6%)≤3 months, 17 studies (33.3%)>6 months.

Among the 51 included RCTs, Five major mHealth intervention types were involved, including, 1) mobile phone text massage (MPTM), 2) mobile phone calls (MPCs), 3) wearable or portable monitoring devices (WPMDs)^[5]^, 4) mobile health applications (mHealth APPs), 5) Telemedicine. The categories follow the principle of simplicity, the ease of intervention, and the degree of public understanding (refer to **Appendix –Table 1**)

### The primary outcome of intervention HbA1c

Forty studies representing 8006 participants reported data on HbA1c, and were pooled for a meta-analysis using the random-effects model. The results showed there were significant mean differences in favour of mHealth interventions compared with traditional treatments for HbA1c control [WMD (95%Cl)=−0.39 (−0.50, −0.29)] (**Figure 2 HbA1c-A**), however, the results demonstrated moderate heterogeneity (*I*^2^ =62.7%, *P*=0.000), so sensitivity analysis by eliminating one study at a time was conducted (**Figure 3 HbA1c-A**). The mHealth interventions had positive impacts in both developed countries [WMD(95%Cl)=−0.35 (−0.46, −0.24)] and developing countries [WMD (95%Cl)=−0.52 (−0.78, −0.26)] (**Figure 2 HbA1c-B**). Furthermore, the subgroup analysis showed that mHealth interventions had a positive effect on all types of diabetes in HbA1c control, and were more significantly in people with T2DM [WMD (95%Cl)=−0.40 (−0.52, −0.28)] than people with T1DM [WMD (95%Cl)=−0.30 (−0.47, −0.12)] (**Figure 2 HbA1c-C**).

**Figure 2.**
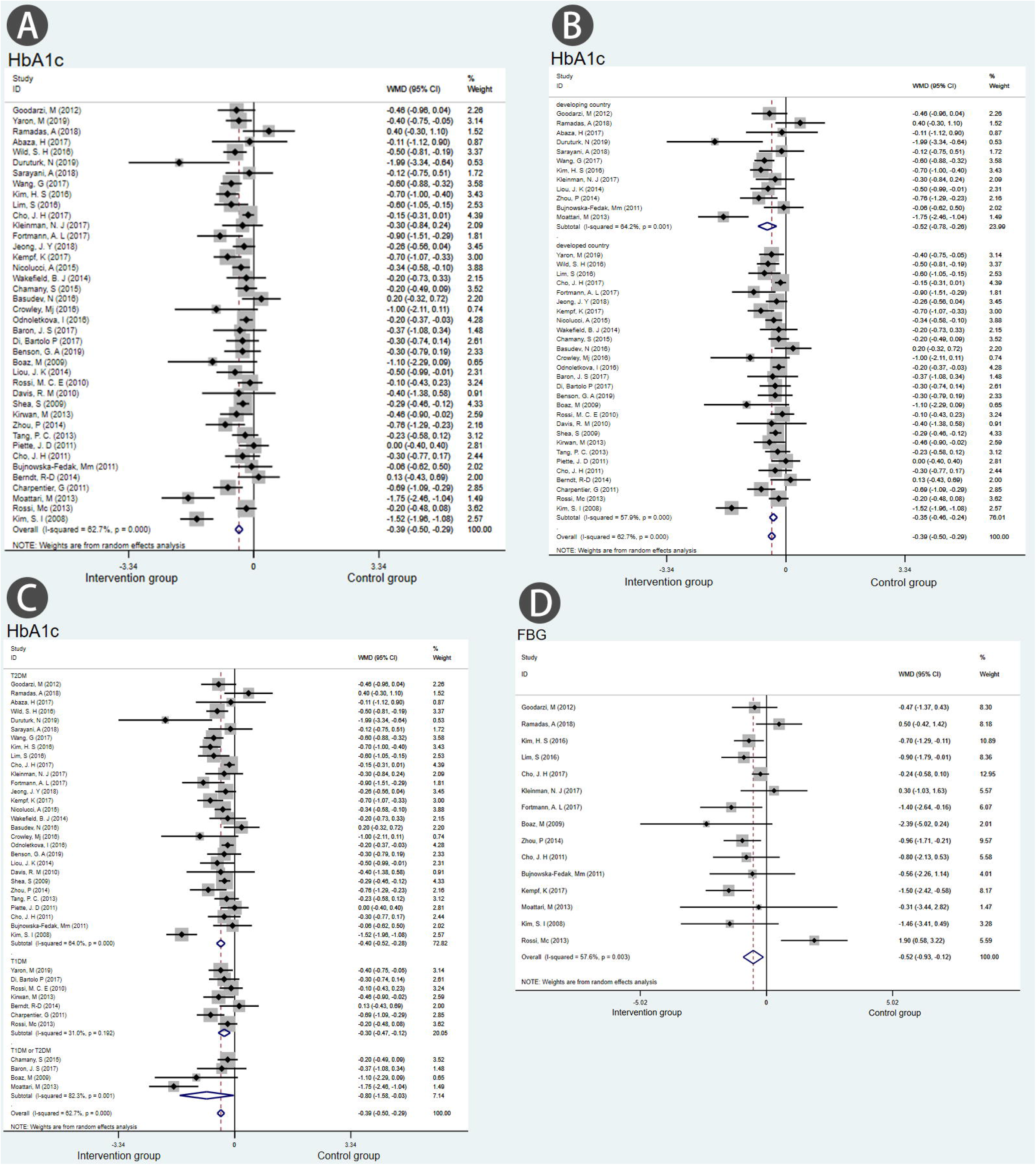
Meta-analyses of mHealth intervention treatments versus other traditional treatments, comparing HbA1c and FBG. Outcomes assessed are (A) change in HbA1c at the end of intervention in studies that compared mHealth treatment with traditional treatment, (B) comparing the effects of mHealth interventions on HbA1c control in Countries with different levels of economic development, (C) comparing the effects of mHealth interventions on HbA1c control in patients with different types of diabetes, and (D) change in FBG at the end of intervention that compared mHealth treatment with traditional treatment.

**Figure 3.**
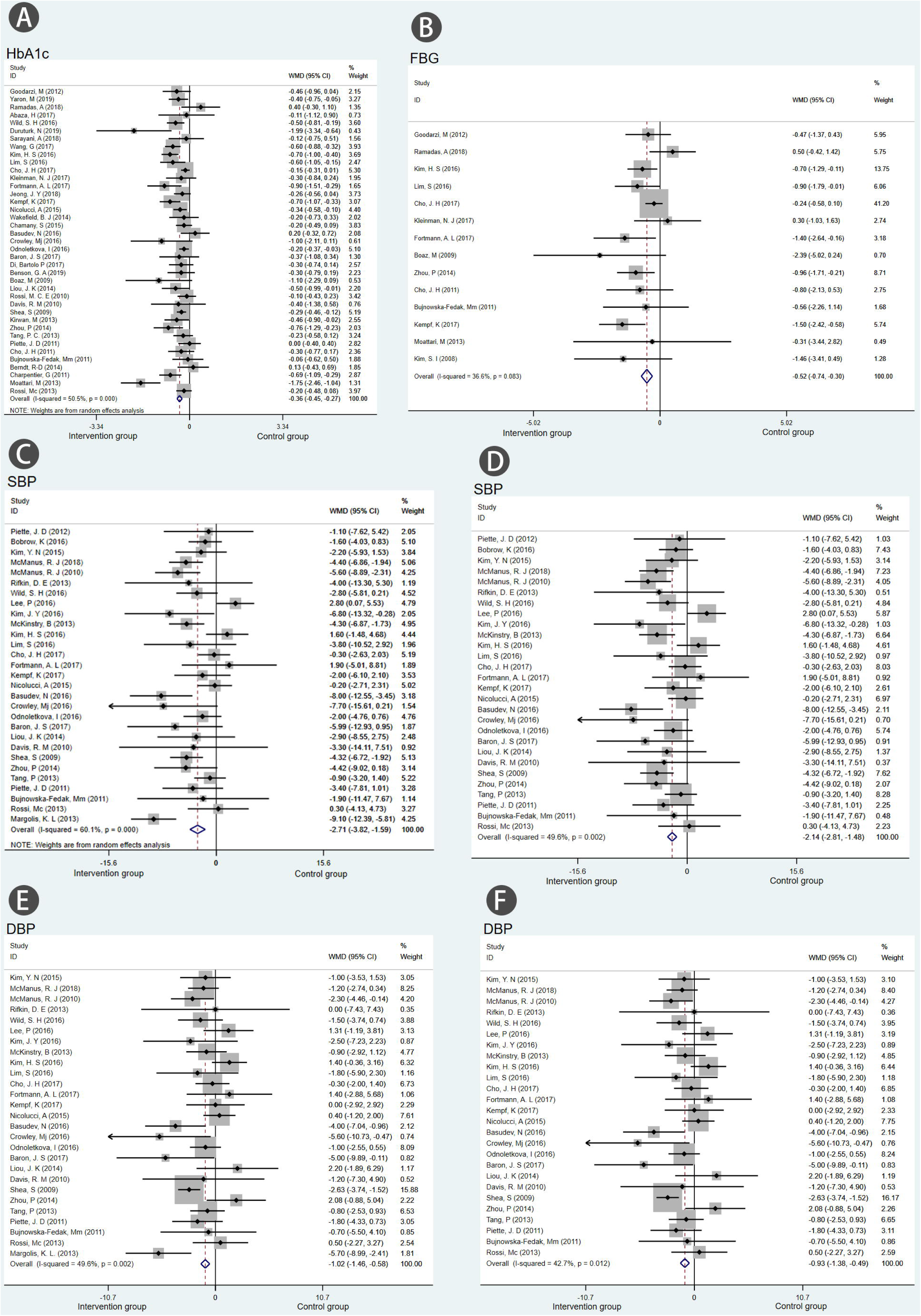
Meta-analyses of mHealth intervention treatments versus other traditional treatments, comparing SBP and DBP. Outcomes assessed are (A) change in SBP at the end of intervention in studies that compared mHealth treatment with traditional treatment, (B) comparing the effects of mHealth intervention on SBP control in Countries with different levels of economic development, (C) SBP in studies that compared combination treatment with mHealth treatment alone, and (D) change in DBP at the end of intervention that compared mHealth treatment with traditional treatment.

In this analysis, there was publication bias on Egger test (*p*=0·036). then, further analysis with trim-and-fill test indicated that the estimates were not affected by publication bias (ie, no trimming performed because data unchanged).

### FBG

In a pooled analysis of 15 trials using the random-effects model, the mHealth interventions led to a mean greater reduction in FBG [WMD (95%CI) =−0.52 (−0.93, −0.12)] **(Figure 2 FBG-D**) than with any other traditional treatment strategy. The results demonstrated moderate heterogeneity (*I*^*2*^=57.6%, *P*=0.003). A sensitivity analysis was conducted **(Figure 3 FBG-B)**. In this analysis, no publication bias was evident (*p*=0·16).

### SBP

In a pooled analysis of 30 trials representing 9476 participants (HTN and DM) reported data on SBP, the mHealth intervention led to a mean higher reduction in SBP [WMD (95%CI) =−2.99 (−4.19, −1.80)] **(Figure 4 SBP-A)** than with any other traditional treatment. But the results demonstrated considerable heterogeneity (*I*^2^=67.3%, *P*=0.000), therefore, we conducted a sensitivity analysis. when Green et al. 2008 was excluded, the *I*^2^ was 60.1%, when Margolis et al. 2013 was excluded, the *I*^2^ was 49.6% **(Figure 3 SBP-C, D)**. There was no significant publication bias in this analysis (*p*=0·439). we conducted a subgroup analysis in countries of different economic levels using mHealth interventions, 11 studies representing 4189 hypertensive patients reported data on SBP, and were pooled for a meta-analysis using the random-effects model. The results showed positive outcomes in favour of mHealth intervention in developed countries [WMD (95%Cl)=−5.72 (−7.46, −3.99)], but no significant difference in developing countries [WMD (95% Cl)=0.25 (−3.10, 3.59)]. In the study, it is reassuring that we found mHealth interventions combined with professional managements are more effective than mHealth interventions alone [WMD (95%Cl)=−6.17 (−8.83, −3.50)] VS [WMD (95%Cl)=−2.16 (−5.07, 0.75)]

### (Figure 4 SBP-B, C). DBP

In a pooled analysis of 28 trials representing 8506 participants reported data on DBP, the mHealth intervention led to a mean greater reduction in DBP [WMD (95%CI) =−1.14 (−1.86, −0.42)] **(Figure 4 DBP-D)** than with any other traditional treatment. Due to the moderate heterogeneity (*I*^2^=57.1%,*P*=0.000), we performed a sensitivity analysis, when Green et al. 2008 and Margolis et al. 2013 were excluded separately, the *I*^2^ was 42.7% **(Figure 3 DBP-E, F)**. In this analysis, no publication bias was evident (*p*=0.857).

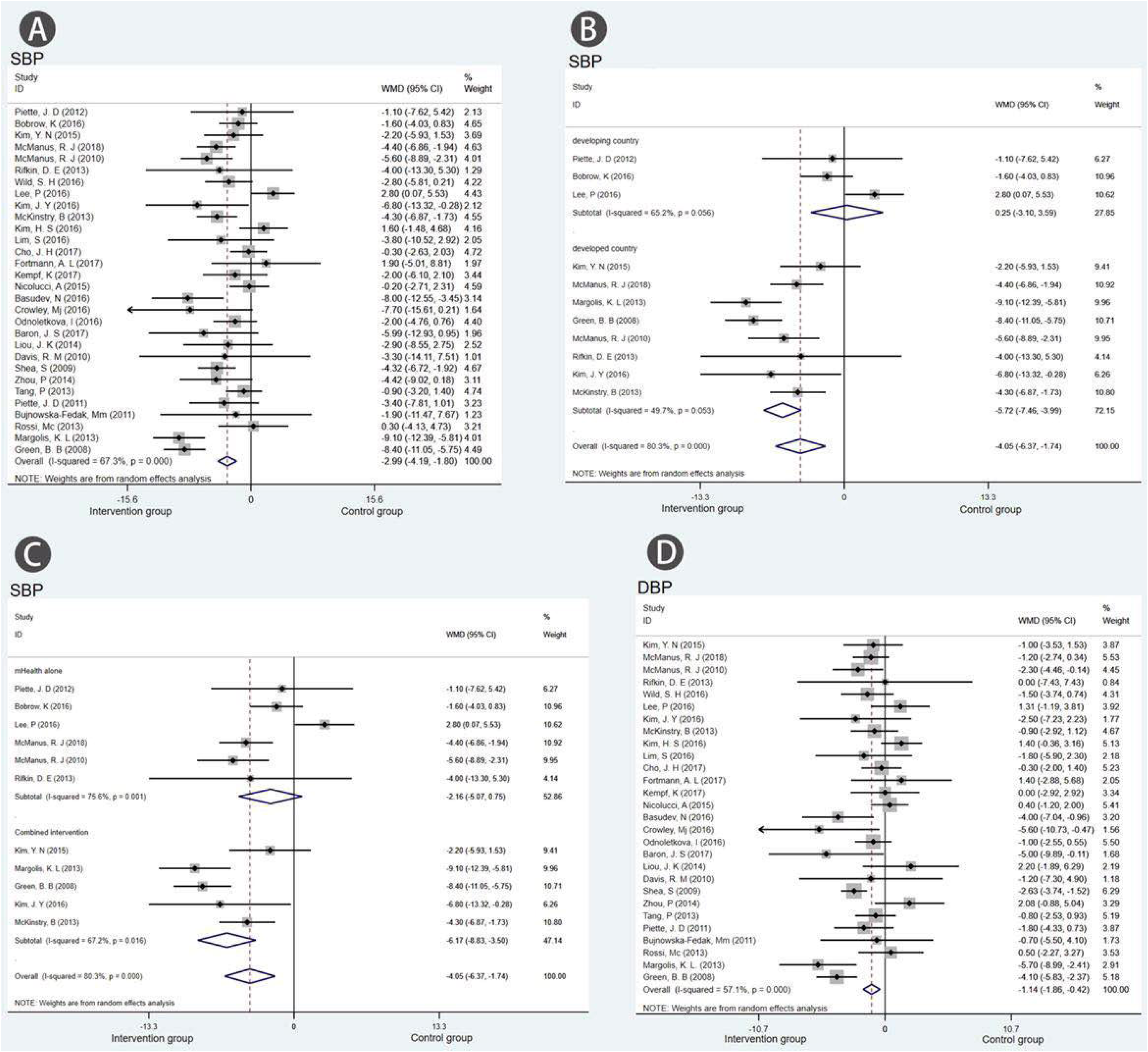

### The secondary results of intervention

**Most o**f all the studies included described positive results, showing that most patients’ clinical indicators improved after using mHealth interventions. 14 studies (27.45%) described the improvement of compliance, 13 (25.49%) reported the improvement of self-care ability, 12 (23.53%) reported the improvement of eating habits and physical exercise, 10 (19.61%) described the change of positive lifestyle, and so on **(Table 3)**.

**Table 3.**
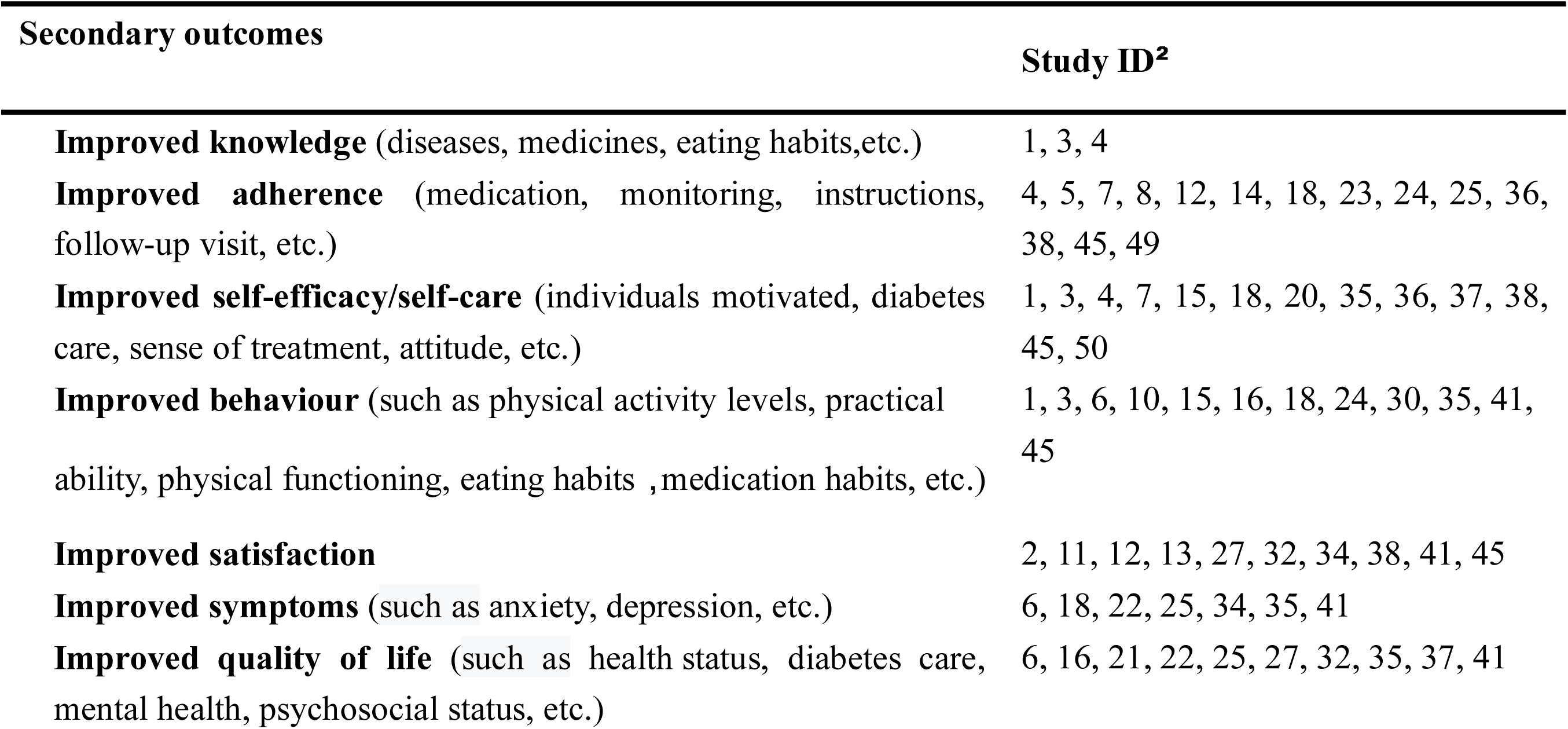
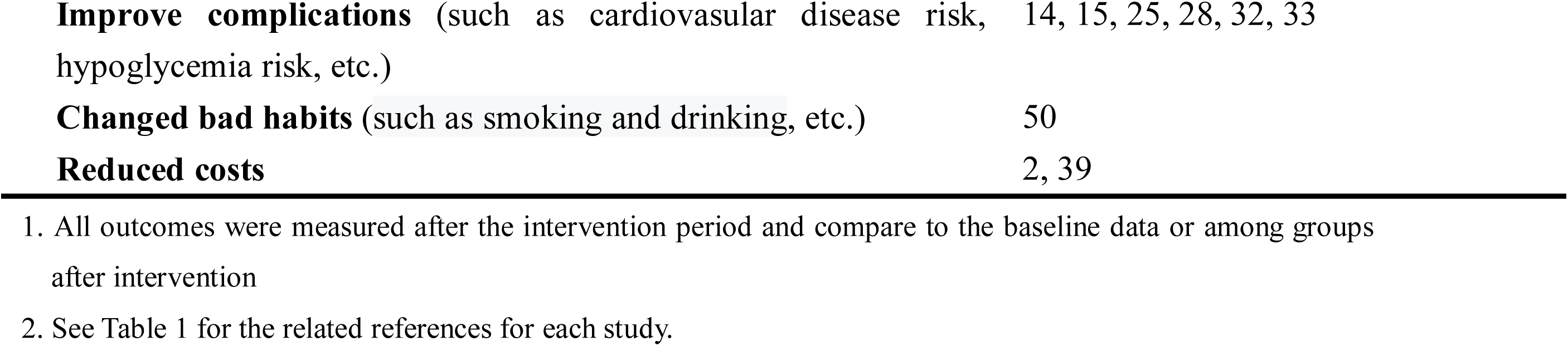
The effects of mHealth interventions on secondary outcomes related to hypertension and diabetes^1^.

## Discussion

This is the first systematic review and meta-analysis to compare the effects of mHealth interventions in countries with different economic development levels, and evaluate the control of clinical outcomes and benefits affter interventions. 51 studies meeting the inclusion criteria were included. Our results show that compared with traditional care treatments, mHealth interventions can yield improved clinical outcomes in HbA1c, FBG, SBP, DBP control in different levels of economic development, and had positive effects on improving quality of life, satisfaction and self-efficacy, etc.

These results enhanced the evidence on the overall effectiveness of mHealth intervention treatments in DM and HTN management as documented in previous studies^[15–17]^ ^[73, 74]^. Most reviews about mHealth interventions before were limited to a single type of intervention, and mainly aimed to evaluate the effect of intervention time and types^[15, 16]^,such as telemedicine^[16] [73]^, mHealth Apps ^[15] [18]^, MPTM^[17]^, etc. However in our review, the mHealth interventions included five types that using in healthcare industries presently. The results we found in our data showed the beneficial effect of mHealth interventions were more pronounced among patients with T2DM than among those with T1DM, which is consistent with Dejun Su,et al’s studies^[16]^. The main reason caused the difference of intervention results may due to the disease itself, as we know patients with T1DM were rely on insulin treatments. But for T2DM, especially in the early stage of diabetes can improve blood glucose by changing lifestyle and eating habits, which was consistent with the direction of mHealth interventions. So how to develop specific interventions for different types of patients are the key to achieving efficacy. Look at the figure 2, we found moderate heterogeneity in the sensitivity analysis of HbA1c, when Kim, S.I et al. 2008 was excluded, the heterogeneity decreased significantly, after checking and comparing the original studies carefully, we inferred that the heterogeneity may be due to the small sample size (N=34).

In our studies, an interesting findings was that the mHealth interventions compare to the control group could significantly improved SBP control in developed countries, but no significant difference in developing countries. after reading, checking and comparing the original studies carefully, we found that the three RCTs (Lee et al. 2016, Piette et al. 2012, Bobrow et al. 2016) performed in the developing countries just using mHealth treatments as intervention alone, didn’t combine the human intelligence which can provide professional guidance about medication, lifestyle, behaviour, etc. But in developed countries the mHealth care usually combined with specialists and professionals to provide disease-related management during the intervention. These results enhanced the evidence on Can Hou et al’s studies ^[15]^ that health care professionals’ functionality is important to achieve clinical effectiveness. To be sure, the BP outcomes in the three RCTs all have positive improvement compare to the baseline after interventions. In our study, we found moderate heterogeneity in the sensitivity analysis of SBP and DBP, when Green et al 2008, Margolis et al 2013 were excluded separately, the heterogeneity decreased significantly. After a detailed analysis of the original study, we found that both articles all had professional pharmacists involving in disease management. we inferred that professionals’ interventions can strengthen management that may be the source of heterogeneity. In order to test the conjecture, we conducted a subgroup analysis to campare the combined intervention with mHealth intervention alone. the results are encouraging, mHealth intervention treatments combined with special staff management (pharmacist, dietitian, specialist nurse and sports trainer) had more effective than mHealth interventions alone [WMD (95%Cl)=−6.17 (−8.83, −3.50)] VS [WMD (95%Cl)=−2.16 (−5.07, 0.75)], The results further validate the above discussion that professionals’ functionality is important to achieve clinical effectiveness.

Nowdays, many reviews ^[15, 16]^ focus on analyzing the effect of mHealth interventions on chronic disease management in developed countries, but lacking the assessment in developing countries ^[5]^. In our studies we included 15 RCTs which conducted in developing countries in recent years, aimed to assess the effectiveness of mHealth interventions in less developed countries. It is reassuring that our review found mHealth could improve the management of chronic diseases in countries with different economic levels, and emphasized that mHealth intervention combined with professionals’ functionality were important to achieve clinical effectiveness.

### Quality of included studies

We evaluated the quality of 51 included studies based on jaded scores, allocation sequences were randomly generated in all trials. Among them, 37 studies (72.55%) reported the concealment of the allocation and addressed incomplete outcome data adequately. This is an open study, given the nature of the intervention, it was not possible to blind patients or their clinicians to their experimental assignment, So double blindness is not feasible. 44 studies (86.27%) described Follow-up reporting, and descriped the reason of dropping out (Appendix-Table 2)

### Limitations

Confounding factors may significantly impact our findings. For example, when we conducted the subgroup analysis by diabetes type of intervention, we did not control for potential differences in baseline HbA1c across the subgroups, futures studies need to explore the findings. However, despite the growing interest in the use of various mobile health technologies, the long-term effects of such interventions are unknown and will need to be tested in a longer and more representative population.

## Conclusion

The present systematic review and meta-analysis indicates that mHealth intervention treatments can improve clinical outcomes, decrease depressive symptoms, improve the quality of life and enhance self-efficacy among patients in countries at all levels of economic development. and emphasized the importance that combined intervention is important to achieve clinical effectiveness.

## Data Availability

The data used to support the findings of this study are included within the article

## Author contributions

All authors have agreed on the final version and meet at least one of the following criteria:1, Substantial contributions to design, acquisition of data or analysis and interpretation of data, 2, Drafting the article or revising it critically for important intellectual content.

## Appendix

**Table 1.**
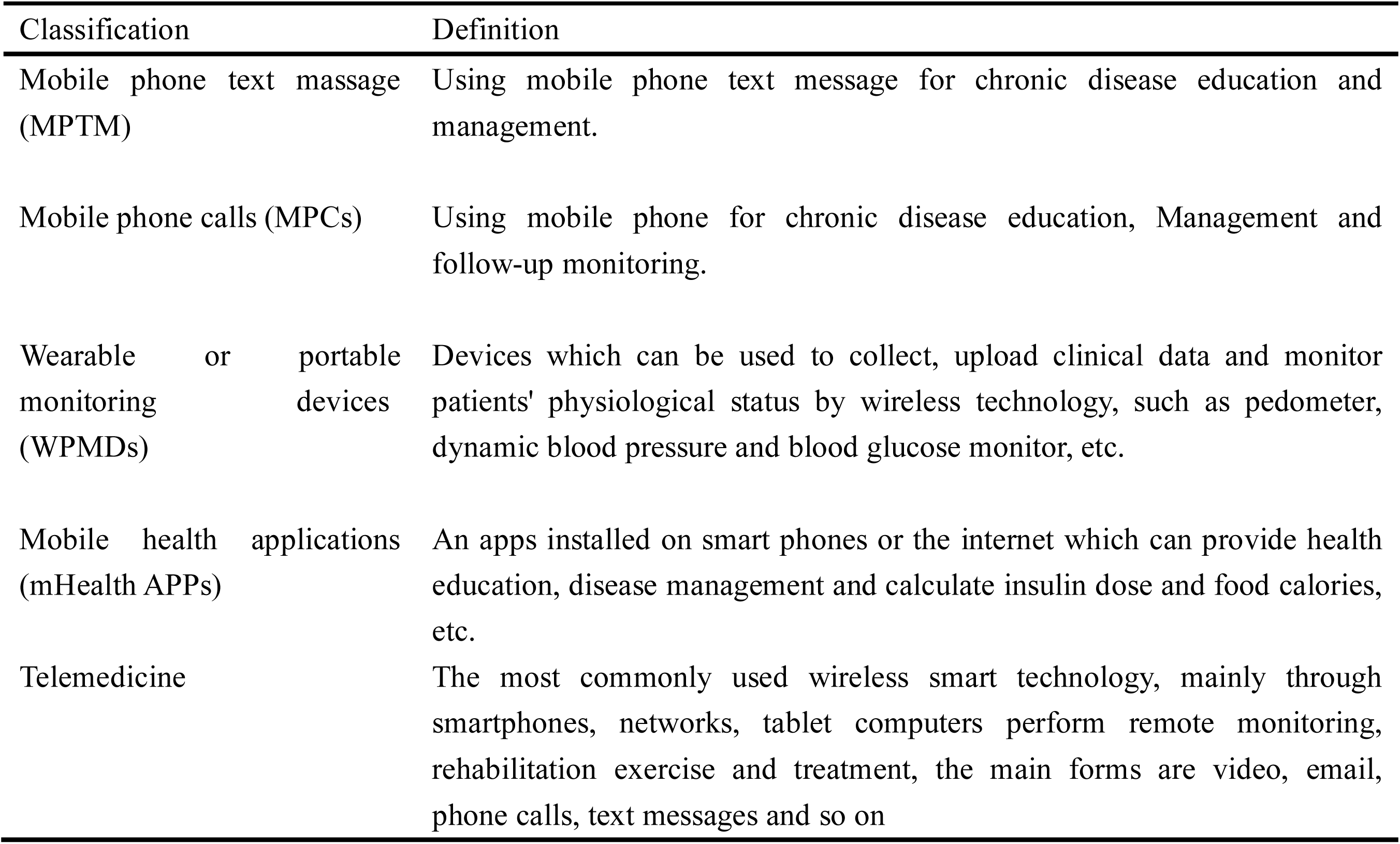
The definition and classification of mobile health in our studies. Classification Definition

| Classification | Definition |
| --- | --- |
| Mobile phone text message (MPTM) | Using mobile phone text message for chronic disease education and management. |
| Mobile phone calls (MPCs) | Using mobile phone for chronic disease education, Management and follow-up monitoring. |
| Wearable or portable monitoring devices (WPMDS) | Devices which can be used to collect, upload clinical data and monitor patients' physiological status by wireless technology, such as pedometer, dynamic blood pressure and blood glucose monitor, etc. |
| Mobile health applications (mHealth APPs) | An apps installed on smart phones or the internet which can provide health education, disease management and calculate insulin dose and food calories, etc. |
| Telemedicine | The most commonly used wireless smart technology, mainly through smartphones, networks, tablet computers perform remote monitoring, rehabilitation exercise and treatment, the main forms are video, email, phone calls, text messages and so on |

**Table 2.** Quality of included all studies

| Study ID<sup>1</sup> | Author | Randomisation | Description of randomisation methods | Double blind | Used identical placebo | Follow-up reporting | Total score |
| --- | --- | --- | --- | --- | --- | --- | --- |
| 1 | Goodarzi, M | 1 | 1 | 0 | 0 | 1 | 3 |
| 2 | Yaron, M | 1 | 0 | 0 | 0 | 1 | 2 |
| 3 | Ramadas, A | 1 | 0 | 0 | 0 | 1 | 2 |
| 4 | Abaza, H | 1 | 1 | 0 | 0 | 1 | 3 |
| 5 | Wild, S. H | 1 | 1 | 0 | 0 | 1 | 3 |
| 6 | Duruturk, N | 1 | 1 | 0 | 0 | 1 | 3 |
| 7 | Sarayani, A | 1 | 1 | 0 | 0 | 1 | 3 |
| 8 | Wang, G | 1 | 0 | 0 | 0 | 1 | 2 |
| 9 | Kim, H. S | 1 | 1 | 0 | 0 | 0 | 2 |
| 10 | Lim, S | 1 | 1 | 0 | 0 | 1 | 3 |
| 11 | Cho, J. H | 1 | 1 | 0 | 0 | 0 | 2 |
| 12 | Kleinman, N. J | 1 | 1 | 0 | 0 | 1 | 3 |
| 13 | Fortmann, A. L | 1 | 1 | 0 | 0 | 1 | 3 |
| 14 | Jeong, J. Y | 1 | 0 | 0 | 0 | 1 | 2 |
| 15 | Kempf, K | 1 | 1 | 0 | 0 | 1 | 3 |
| 16 | Nicolucci, A | 1 | 1 | 0 | 0 | 1 | 3 |
| 17 | Wakefield, B. J | 1 | 1 | 0 | 0 | 1 | 3 |
| 18 | Chamany, S | 1 | 1 | 0 | 0 | 1 | 3 |
| 19 | Basudev, N | 1 | 1 | 0 | 0 | 1 | 3 |
| 20 | Rossi, M. C. E | 1 | 1 | 0 | 0 | 1 | 3 |
| 21 | Odnoletkova, I | 1 | 1 | 0 | 0 | 1 | 3 |
| 22 | Baron, J. S | 1 | 1 | 0 | 0 | 1 | 3 |
| 23 | Di, Bartolo P | 1 | 0 | 0 | 0 | 1 | 2 |
| 24 | Benson, G. A | 1 | 0 | 0 | 0 | 1 | 2 |
| 25 | Boaz, M | 1 | 0 | 0 | 0 | 1 | 2 |
| 26 | Liou, J. K | 1 | 0 | 0 | 0 | 1 | 2 |
| 27 | Rossi, M. C. E | 1 | 1 | 0 | 0 | 1 | 3 |
| 28 | Davis, R. M | 1 | 0 | 0 | 0 | 0 | 1 |
| 29 | Shea, S | 1 | 0 | 0 | 0 | 1 | 2 |
| 30 | Kirwan, M | 1 | 1 | 0 | 0 | 1 | 3 |
| 31 | Moattari, M | 1 | 1 | 0 | 0 | 1 | 3 |
| 32 | Rossi, Mc | 1 | 1 | 0 | 0 | 1 | 3 |
| 33 | Zhou, P | 1 | 1 | 0 | 0 | 1 | 3 |
| 34 | Tang, P | 1 | 1 | 0 | 0 | 1 | 3 |
| 35 | Piette, J. D | 1 | 1 | 0 | 0 | 0 | 2 |
| 36 | Cho, J. H | 1 | 1 | 0 | 0 | 1 | 3 |
| 37 | Bujnowska-Fedak, Mm | 1 | 0 | 0 | 0 | 1 | 2 |
| 38 | Berndt, R-D | 1 | 0 | 0 | 0 | 0 | 2 |
| 39 | Charpentier, G | 1 | 1 | 0 | 0 | 1 | 3 |
| 40 | Kim, S. I | 1 | 1 | 0 | 0 | 1 | 3 |
| 41 | Piette, J. D | 1 | 1 | 0 | 0 | 0 | 2 |
| 42 | Bobrow, K | 1 | 1 | 0 | 0 | 1 | 3 |
| 43 | Kim, Y. N | 1 | 1 | 0 | 0 | 1 | 3 |
| 44 | McManus, R. J | 1 | 1 | 0 | 0 | 1 | 3 |
| 45 | Margolis, K. L | 1 | 0 | 0 | 0 | 1 | 2 |
| 46 | Green, B. B | 1 | 1 | 0 | 0 | 1 | 3 |
| 47 | McManus, R. J | 1 | 1 | 0 | 0 | 1 | 3 |
| 48 | Rifkin, D. E | 1 | 1 | 0 | 0 | 1 | 3 |
| 49 | Lee, P | 1 | 1 | 0 | 0 | 1 | 3 |
| 50 | Kim, J. Y | 1 | 0 | 0 | 0 | 0 | 1 |
| 51 | McKinstry, B | 1 | 1 | 0 | 0 | 1 | 3 |

<sup>1</sup>Study IDs indicate the 1st to 51th study. Jadad scores range from 0 (very poor) to 5 (rigorous) and consist of points randomization (randomized=1 point; table of random numbers or computer-generated randomization=additional 1 point), double blindness (double blind=1 point; use masking such as identical placebo=additional 1 point), and follow-up (stating numbers of subjects withdrawn and the reasons for them in each group of a study=1 point).

